# ClarID: A Human-Readable and Compact Identifier Specification for Biomedical Metadata Integration

**DOI:** 10.1101/2025.09.05.25335150

**Authors:** Manuel Rueda, Ivo G. Gut

**Affiliations:** Centro Nacional de Análisis Genómico, C/Baldiri Reixac 4, 08028 Barcelona, Spain; Universitat de Barcelona (UB), Barcelona, Spain

**Keywords:** Health data integration, Subject identifiers, Biosample identifiers, Structured Barcoding, Metadata harmonization, Ontology-based annotation

## Abstract

**Background:** In biomedical research, subjects and biospecimens are commonly tracked using simple IDs or UUIDs, which guarantee uniqueness but convey no embedded semantic information. Contextual metadata (such as tissue type, diagnosis, or assay) is often stored separately, making integration, cohort selection, and downstream analysis cumbersome. While structured barcoding systems exist in large consortia (e.g., TCGA, GTEx) or domain-specific contexts (e.g., SPREC, GOLD), no unified, extensible framework currently spans both subjects and biosamples in a human- and machine-readable way.

**Methods:** We developed ClarID, a domain-agnostic specification that supports two identifier formats: (i) a human-readable form (e.g., ‘CNAG_Test-HomSap-00001-LIV-TUM-RNA-C22.0-TRT-P1W’ that encodes key metadata such as project, species, subject_id, tissue, assay, disease, timepoint and duration (from that event); and (ii) a compact version named ‘stub’ (e.g., ‘CT01001LTR0N401T1W’) optimized for filenames, pipelines, and labeling.

ClarID is implemented through an open-source command-line tool, ClarID-Tools, which processes tabular metadata files (CSV/TSV) and uses a YAML-based codebook to generate, decode, and validate identifiers, as well as to create and read QR codes. The tool supports bulk and single-sample processing and allows easy integration with institutional workflows.

**Results:** To demonstrate ClarID’s utility, we applied it to datasets from the Genomic Data Commons (GDC), generating interpretable identifiers for more than 113,000 clinical records (subjects) and 4,255 biospecimen records. All materials, including pre-processing scripts, input and encoded data, are publicly available and fully reproducible via the accompanying GitHub repository and Google Colab.

**Conclusions:** ClarID fills a critical gap between opaque accession numbers and rich metadata schemas by embedding key context directly into structured identifiers. It enhances traceability, facilitates downstream analysis, and remains adaptable to project-specific needs through a configurable codebook. The accompanying ClarID-Tools software is freely available, together with full documentation and reproducible pipelines, at https://github.com/CNAG-Biomedical-Informatics/clarid-tools.

## BACKGROUND

Reliable and unambiguous identification of research subjects and biospecimens is a foundational requirement in biomedical research (1–3). Most current systems rely on programmatically generated identifiers—such as incrementing numbers, alphanumeric strings, or universally unique identifiers (UUIDs)—which guarantee uniqueness but typically lack semantic meaning (4). For example, a UUID such as ‘550e8400-e29b-41d4-a716-446655440000’ offers no immediate information about the species, tissue type, or disease context it represents. This lack of embedded meaning hinders data exploration and slows downstream analysis, as researchers must rely on external metadata files even for basic contextual information.

To improve sample traceability and metadata standardization, several domain-specific solutions have emerged (see Table 1). For instance, ISBT 128, an international standard for labeling medical products of human origin, supports robust biospecimen tracking through globally unique identifiers such as Donation Identification Numbers (DINs), standardized product codes, and encoded 2D barcodes (5). By including information such as collection date, specimen type, and aliquot details, ISBT 128 supports traceability and regulatory compliance in both clinical and research contexts. Complementing this, the BRISQ (Biospecimen Reporting for Improved Study Quality) guidelines aim to standardize the reporting of preanalytical variables, such as collection method and storage conditions, to improve data transparency and reproducibility (6). While BRISQ is not a barcoding system per se, it underscores the importance of structured metadata capture.

**Table 1.** Example Identifiers Used in Major Coding Systems.

| System | Example Identifier | Format Notes |
| --- | --- | --- |
| ClarID (human) | CNAG_Test-HomSap-00001-LIV-TUM-RNA-C22.0-TRT-P1W-B01-R05 | Verbose, human-readable identifier with hyphen-separated components: project, species, subject_id, tissue, sample type, assay, condition(s), timepoint, duration, batch (opt.), replicate (opt.) |
| ClarID (stub) | CT01001LTR0N401T1WB01R05 | Compact identifier using abbreviated codes |
| TCGA | TCGA-02-0001-01C-01D-0182-06 | 9-field barcode encoding tissue source site, analyte, center, etc. |
| GTEX | GTEX-14753-1626-SM-5NQ9L | 5 parts: project, donor ID, tissue code, sample marker, aliquot |
| SPREC | TIS-SPS-N-E3-FPL-M-CQ** | 7-element code for preanalytical variables for fluid/solid samples (e.g., sample type, container, storage) |
| GOLD | Gb0123456 | Accession-style ID (Gb:biospecimen); metadata stored separately in database |
| ISBT 128 | R1234 25 987654 | DIN (Donation ID Number); globally unique, traceable |
| Schrade et al. | ACCAndDivT00001 | 17-char code: taxonomy + sample type + 5-digit ID |
| Kurtz et al. | UKBi001-A-1 | Institution + donor + cell type + clone ID (+ subclone) |

\*\* Fluid sample

Several large-scale genomics initiatives have embedded hierarchical metadata directly into structured identifiers. The Cancer Genome Atlas (TCGA) (7) developed a nine-field hierarchical barcoding system encoding project, tissue source site, participant, sample type, and analytical details, enabling researchers to infer sample context without consulting external files. Similarly, the Genotype-Tissue Expression (GTEx) consortium (8) employs a five-part barcode structure that captures donor identity and tissue site.

The Sample PREanalytical Code (SPREC), developed by the International Society for Biological and Environmental Repositories (ISBER), takes a different approach by encoding sample type, handling, and storage conditions into a standardized code for biospecimen quality documentation (9). SPREC promotes harmonized practices across biobanks and enhances reproducibility in biomarker research. Similarly, the Genomes OnLine Database (GOLD) addresses inconsistencies in microbiome sample naming by embedding environmental, biological, and geographic metadata into structured identifiers (10). This system has been applied to over 180,000 samples, underscoring the importance of contextual naming for comparative metagenomic analyses.

In the field of wildlife biology, Schrade et al. proposed a taxonomy-aware, human-readable identifier format for biospecimens, integrating species and provenance data into a 17-character barcode (11). Their system combines taxonomic abbreviations, species binomials, sample type codes, and serial numbers (e.g., ACCAndDivT00001) and advocates for a centralized registry to promote long-term stability and FAIR principles (12). Similar efforts have emerged for pluripotent stem cell lines, such as the system developed by Kurtz et al., which supports traceability of donor-derived lines through consistent nomenclature (13).

Despite the range of existing proposals, the adoption of structured identifiers or barcoding systems remains limited. Several factors contribute to this: some systems are tightly coupled to specific projects—such as TCGA—while others, like SPREC, are designed primarily for biospecimen labeling and traceability in clinical biobanking. A significant barrier is the lack of accessible, programmatic tooling; the burden of generating structured identifiers is often left to individual users, with little to no software implementations, limiting scalability and broader adoption.

Moreover, aside from the TCGA barcoding system that embeds subject id on the barcode, no standardized framework currently exists (to our knowledge) for assigning structured identifiers at the subject (individual) level. In clinical studies, a single subject may be linked to multiple biospecimens, yet the associated phenotypic or diagnostic metadata is often detached from the identifiers. This forces researchers to rely on flat CSV files or manual dataset merges, a common workflow in repositories such as dbGaP and EGA. That approach adds friction and increases the risk of error in everyday tasks such as quickly identifying metadata from a subject or sample ID..

To address these challenges, we introduce ClarID, a lightweight identifier specification that supports both subject- and biosample-level encoding. ClarID provides a project- and disease-agnostic framework that embeds essential metadata directly into the identifier, resulting in self-documenting labels that are readable, semantically transparent, and easy to interpret. Inspired by structured schemes such as TCGA, GTEx, SPREC, and the system by Schrade et al., ClarID encodes each identifier as a sequence of components, each drawn from controlled vocabularies defined in a configurable YAML codebook. An open-source command-line tool (ClarID-Tools) enables bulk encoding, decoding, validation, and QR (quick-response) code generation. Identifiers can be generated in a verbose, human-readable format or in a condensed alternative called the Simple Terse Universal Barcode (STUB). The stub format prioritizes compactness while retaining limited interpretability, similar to how CIGAR strings in SAMtools (14) concisely encode alignment operations for downstream use. ClarID does not aim to replace formal metadata standards, but instead offers a practical middle ground: unique, ontology-linked identifiers that embed essential metadata for rapid contextual understanding, without the need to query external files. By bridging human readability with machine-actionable semantics, ClarID provides a scalable solution for subject and sample identification across biomedical research domains.

In the sections that follow, we describe the conceptual framework behind ClarID, demonstrate its practical application through a real-world datasets from GDC and compare it to existing barcode and naming standards.

## METHODS

### Overview

The core encoding and decoding processes are handled by ClarID-Tools software, which serves as the reference implementation for the ClarID specification. This system is powered by a YAML-based codebook (structured dictionary in text format) that defines the vocabulary for each domain-specific attribute (see Figure 1). While the ClarID-Tools software will be detailed later, we begin here by describing the structure and logic of the identifiers themselves.

**Figure 1:**
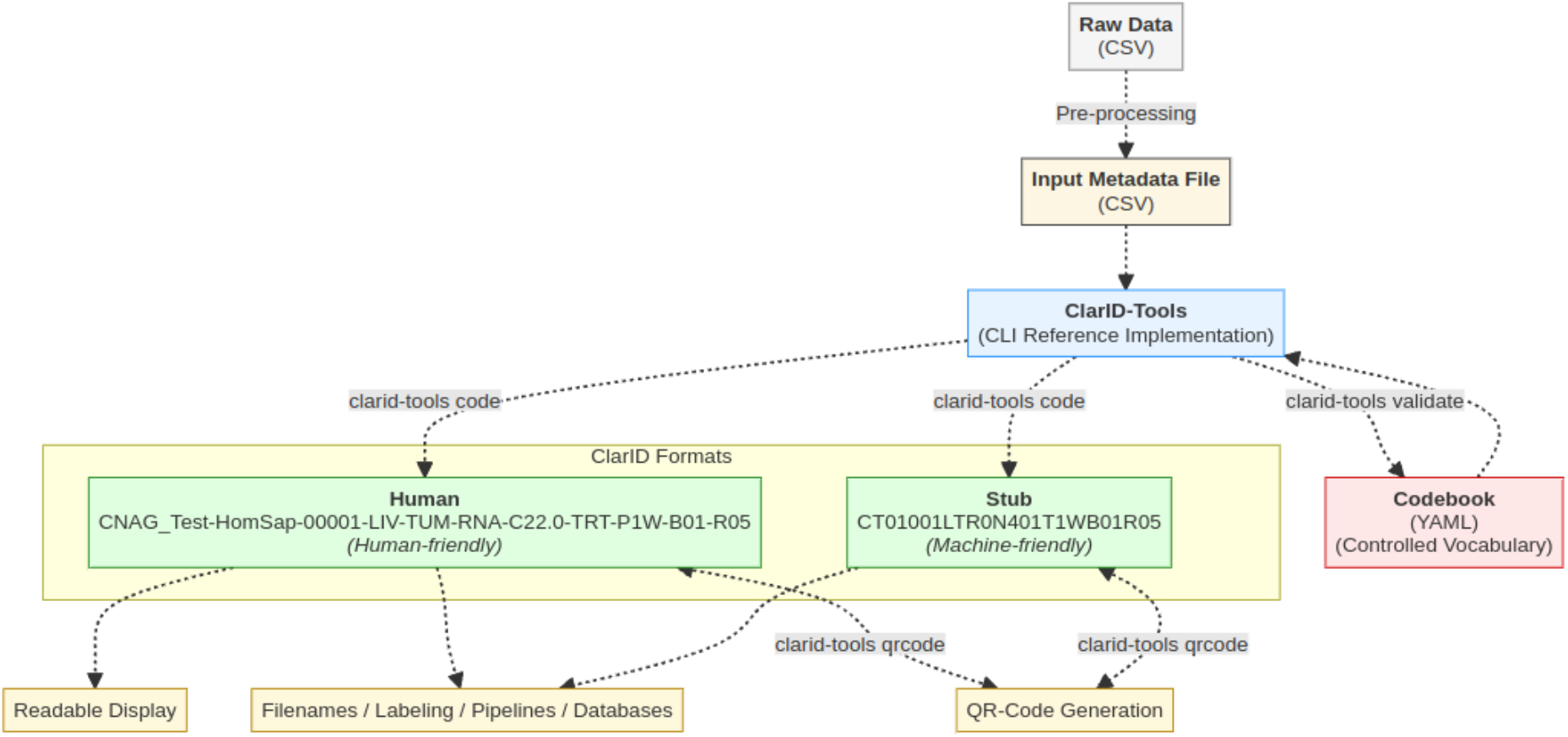
ClarID generation workflow. Raw CSV data is pre-processed into structured metadata and processed by the ClarID-Tools CLI using a controlled YAML codebook. The tool produces both human-readable and compact stub ClarIDs, which can be encoded or decoded, and are used for labeling, display, databases, and QR code generation.

The need for structured identifiers emerged during our efforts to match patients in large cohorth, such as those from the GDC, using our Pheno-Ranker tool (15). To inform our approach, we conducted a comprehensive review of existing conventions in established repositories (see Table 1). While we drew inspiration from prior formats such as the TCGA barcode and the schema proposed by Schrade et al., it became clear that a more flexible solution was needed—one balancing semantic clarity with compactness. In designing ClarID, we prioritized practical requirements for downstream analysis as well as ease of interpretation by human users. This led us to develop two complementary formats: the human-readable format (“human”) and the compact format (“stub”). The stub format is inspired in part by conventions like the CIGAR string format used in SAMtools (14), offering concise, information-dense codes suited for file and database usage.

### Identifier Generation and Formats

The system supports encoding (ID creation) and decoding (ID interpretation) for two types of entities: “biosample” (also referred to as “biospecimen”, with the terms used interchangeably herein) and “subject”. Each identifier is composed of a sequence of categorical values, arranged in a predefined order and referred to as components, each drawn from a controlled vocabulary defined in the codebook.

In *human* format (see Table 2), identifiers are verbose and semantically informative. For biosamples, an identifier consists of nine mandatory fields (<project.code>-<species.code>-<subject_id>-<tissue.code>-<sample_type.code>-<assay.code>-<condition>-<timepoint.code>-<duration.code>) and two optional ones (<batch>-<replicate>) all separated by hyphens. A typical example is ‘CNAG_Test-HomSap-00001-LIV-TUM-RNA-C22.0-TRT-P1W-B01-R05’. Each component encodes a specific attribute: project (CNAG_Test), species (HomSap for *Homo sapiens*), zero-padded subject ID (00001), tissue (LIV, liver), sample type (TUM, tumor), assay (RNA, RNA-seq), clinical condition (ICD-10 code C22.0, liver cell carcinoma), timepoint (TRT, treatment event), duration relative to that event (P1W, 1 week, ISO 8601 format, max. 3 chars), batch (B01), and replicate (R05) (see Table 2). For subjects, an identifier consists of six mandatory fields separated by hyphens (<study.code>-<subject_id>-<type.code>-<condition.code>-<sex.code>-<age_group.code>). A typical example is ‘COPDStudy-01001-Case-J44.9-Male-A40_49’. Each component represents: study name (COPDStudy), zero-padded subject ID (01001), type (Case), clinical condition (ICD-10 code J44.9), sex (Male), and age group (A40_49) (see Table 2). In the human-readable format, most components are defined by their content rather than by a fixed number of characters, though some fields (e.g., zero-padded IDs or codes) do have a defined length.

**Table 2.** ClarID Identifier Formats for Subjects and Biospecimens in Human-Readable and Stub Formats.

| Entity | Format | Format Syntax | Example | Description |
| --- | --- | --- | --- | --- |
| Biosample | Human | Project-species-subject_id-tissue-sample_type-assay-condition-timepoint-batch-replicate | CNAG_Test-HomSap-00001-LIV-TUM-RNA-C22.0-TRT-P1W-B01-R05 | CNAG Test project sample, *Homo sapiens*, ID 1, liver, tumor, RNA-seq, ICD10: C22.0, Treatment, Week 1, batch 1, replicate 05 |
| Biosample | Stub | Project-species-subject_id-tissue-sample_type-assay-condition-timepoint-batch-replicate | CT01001LTR0N401T1WB01R05 | Compact identifier version encoding the same metadata as above |
| Subject | Human | study-subject_id-type-condition-sex-age_group | COPDStudy-01001-Case-J44.9-Male-A40_49' | Subject in COPD study, ID 1001, Case, ICD10: J44.9, male, aged 40–49 |
| Subject | Stub | study-subject_id-type-condition-sex-age_group | COPDStudy0G9C3Of01MA4 | Compact subject identifier with same metadata as above |

In *stub* format, identifiers are compact, omitting delimiters and using short codes. For instance, the biosample ID ‘CT01001LTR0N401T1WB01R05’ encodes the same information as above, using stub codes from the codebook as well as internal transformations. Similarly, a subject stub id like ‘COPDStudy0G9C3Of01MA4’ condenses the same elements into a shorter string (see more technical details on the encoding logic in Additional File 1).

### Codebook Architecture

The codebook is implemented as a YAML document (YAML Ain’t Markup Language), a widely used, human-readable format for structured data, well suited to configuration files due to its clarity and ease of manual editing. It is organized around the “biosample” and “subject” entities. Each component (e.g., project, species, tissue) is defined as a key, and its subkeys map to both a long-form “code” and a compact-form “stub_code”. For example:

~~~
biosample:
  project:
    “TCGA-AML”:
    code: TCGA_AML stub_code: AML
    label: “TCGA Acute Myeloid Leukemia”
    id: “NCIT:C17998” # Unknown
  species:
    Human:
    code: HomSap stub_code: 01
    label: “Homo sapiens”
    id: “NCBITaxon:9606”
    tax_code: MPH
  tissue:
   Liver:
    code: LIV
    stub_code: L
    label:”Liver”
    id: “UBERON:0002107”
  assay:
   RNA_seq:
    code: RNA
    stub_code: R
    label: “RNA-seq”
    id: “EFO:0008896”
…
~~~

The fields “code” and “stub_code” are strings used for encoding, while “id” and “label” are included in the codebook for reference only. The latter follow the *ontologyClass* structure from the Phenopacket v2 schema (16), where “id” is a CURIE-style identifier (compact prefix:reference, e.g., UBERON:0001690) and “label” is the human-readable name of the class. CURIE identifiers vary by component: for example, the “tissue” field uses terms from UBERON—a cross-species anatomy ontology that semantically integrates anatomical structures across Metazoa (17) and BRENDA tissue ontology (BTO) (18). For the “condition” field (i.e., disease), ICD-10 diagnoses codes are used. Conditions are not included in the main codebook but are handled internally through a bundled JSON file. When decoding, ClarID-Tools can optionally translate ICD-10 codes back into condition names using the *--with-condition-name* flag. This feature relies on an internal lookup table containing ICD-10 codes and definitions. ClarID does not include a dedicated component for analyte (DNA, RNA, protein); instead, it can be inferred from the “assay” field (e.g., RNA-seq implies RNA). Certain properties, such as “duration”, “batch”, or “replicate”, are defined by regular expressions, as enumerating all possible combinations would be impractical.

The “subject” section similarly defines study, subject ID, type, sex, age group using a uniform schema. The “study” field has the same function as the “project” field in biosample identifiers: it is free form by default but can be restricted to a controlled vocabulary in the codebook if desired. See the full description of the codebook in Additional File 1.

### Reference Implementation Details: ClarID-Tools

ClarID-tools is implemented as a command-line interface (CLI), with the current reference version written in Perl 5, designed for simplicity and portability (see Additional File 1). Full implementation details, including usage examples, configuration, and validation logic, are provided in the documentation at https://cnag-biomedical-informatics.github.io/clarid-tools. In Additional File 2: Table ST1, we describe how we addressed key challenges encountered during the development of ClarID-Tools.

The *code* subcommand handles both encoding and decoding of ClarID identifiers. It supports two modes of operation, controlled by the *--action* option: *encode* and *decode*. The format of the resulting IDs (either human-readable or compact) is selected using the *--format* option (*human* or *stub*).

The *validate* subcommand checks that a given codebook YAML file conforms to a specified JSON Schema. When run with debugging enabled, it can also self-validate the schema.

The *qrcode* subcommand can generate and read back both human and stub ClarID identifiers as QR codes (See Additional File 3, Figure SF1).

Together, these subcommands enable reproducible and schema-driven identifier generation, improving traceability and interoperability in biomedical datasets.

## RESULTS

### Use Case I: Subject-Level Encoding of GDC Data

On June 1, 2025, we downloaded clinical metadata from the Genomic Data Commons (GDC) portal as part of the archive ‘clinical.cohort.2025-06-02.tar.gz’, which included four TSV files: ‘clinical.tsv’, ‘family_history.tsv’, ‘follow_up.tsv’, and ‘pathology_detail.tsv’. We focused on ‘clinical.tsv’, which contained 210 columns and 113,760 rows across 86 studies (including 33 from TCGA). Missing or non-reported values were standardized as ‘NA’.

To prepare the data for ClarID encoding, we applied a preprocessing pipeline detailed in the project’s GitHub repository and documented in the accompanying Google Colab notebook. Steps included selecting relevant clinical fields, converting unique_id (UUID) to a numeric “subject_id”, and binning age values from “demographics.age_at_index” to match predefined ranges in the codebook. ClarID mappings were configured externally via a YAML mapping file, allowing the same process to be adapted to other datasets without modifying the conversion script.

The objective of using ClarID was to provide semantic meaning to otherwise opaque identifiers (e.g., UUIDs), making them interpretable briefly. After parsing, we generated two output files: one containing human-readable ClarID codes, and another with the compact stub format. Both were created using the ‘clarid-tools’ CLI utility. An example of ClarID in human format is ‘TCGA_LIHC-00003-Case-C22.0-Male-A40_49’ and the same in stub format TCGA-LIHC003C0N401MA4’. All scripts, input files, configuration, and results are available in the GitHub repository at https://github.com/CNAG-Biomedical-Informatics//clarid-tools and can be reprodudced with the included Google Colab. A subset of the human-format encodings is available for browsing in an interactive table in the documentation at https://cnag-biomedical-informatics.github.io/clarid-tools.

### Use Case II: Biosample-Level Encoding of GDC Data

In parallel with the subject-level analysis, we performed ClarID encoding at the biosample (more specifically, biospecimen) level. For this use case, we selected only one project (TARGET-AML) within the file ‘biospecimen.project-target-aml.2025-06-29.tar.gz’ downloaded from the GDC portal. The unpacked file consisted of four TSV files (‘aliquot.tsv’, ‘analyte.tsv, ‘portion.tsv’, ‘sample.tsv’, ‘slide.tsv’). We used the file ‘sample.tsv’ that contained 4255 rows, each with 39 columns. After performing a series of preprocessing steps, such as column selection, normalization, and formatting, we generated a structured CSV file suitable for ClarID processing.

Using the ‘clarid-tools’ command-line utility, we encoded the biosample data into both the human-readable and compact stub formats. An example of ClarID in human format is TARGET_AML-HomSap-00002-BMR-PRI-NAV-C92.0-COL-P0N, where each component encodes a specific attribute: BMR, bone marrow; PRI, primary tumor site; C92.0, ICD-10 code for acute myeloblastic leukemia (AML); COL, collection time; and P0N, duration not available. The same information, when encoded in stub format, is ‘TAML01002RPn0iC01CT0N’.

As in Use Case I, all resulting files from both use cases are available in the GitHub repository, along with the full conversion pipeline. The preprocessing scripts are configurable for any CSV dataset and are fully documented, with a reproducible workflow available through a Google Colab notebook (see online documentation). In addition, all human-format encodings can be browsed in an interactive table at https://cnag-biomedical-informatics.github.io/clarid-tools.

## DISCUSSION

### Rationale and Design Philosophy

We introduce a new class of identifiers intended to support downstream analysis by providing immediate, interpretable context about subjects and biospecimens. These identifiers are not meant to replace existing unique identifiers (e.g., UUIDs), but rather to serve as an additional variable—human-readable and semantically informative—that offers quick insights without consulting external metadata files such as CSVs. This design was motivated by recurring needs at our institution, where rapid assessment of sample context was often hindered by fragmented metadata.

ClarID leverages our experience in health data harmonization (19,20) within the framework of the GA4GH (Global Alliance for Genomics and Health) standards (21). To support usability, the reference codebook is pre-populated with example values for attributes such as species, tissues, and assays. These are enriched with ontology mappings, while the codebook remains extensible for institution- or domain-specific terms.

To ease integration into existing workflows, we developed ClarID-Tools, an open-source command-line utility for single or batch encoding/decoding and QR code generation of identifiers directly from CSV or TSV files.

### Comparison to Existing Barcodes

Numerous barcoding schemes have been developed to encode biospecimen metadata, often tailored to specific projects or domains (see Table 3). While many are robust within their original context, they typically lack the flexibility or generalizability needed for broader use. ClarID addresses this gap by offering a compact, domain-agnostic identifier framework that supports both subject- and biospecimen-level encoding, enabling rapid, semantically meaningful decoding for downstream research.

**Table 3.** Comparison of Structured Identifier Systems for Biospecimens Across Biomedical Domains.

| System | Metadata Structure | Domain Specificity | Standardization | Tooling & Support | Adoption Scale |
| --- | --- | --- | --- | --- | --- |
| **ClarID (biosample)** | 9 fields (+ 2 optional): project, species, subject_id, tissue, sample_type, assay, condition, timepoint, duration (*optional*: batch, replicate); YAML-based | Domain-agnostic (human/non-human) | Centralized, extensible codebook | Open-source (ClarID-Tools) | Emerging, designed for broad use |
| **TCGA** | 9 fields: project, TSS, participant, sample type, vial, portion, analyte, plate, center | Cancer genomics | Project-specific, fixed format | Internal scripts only (BCR-LIMS) | Moderate (TCGA consortium) |
| **GTEx** | 5 fields: project, donor ID, tissue site code, sample marker, aliquot ID | Human transcriptomics | Semi-standardized; opaque codes | Metadata files + doc lookup | Moderate (GTEx project) |
| **SPREC** | Encodes preanalytical variables (collection, processing, storage) in fixed code | Biobanking & clinical research | ISBER standard | Manual / Spreadsheet-assisted (SPRECalc) / LIMS-integrated (SPRECware) ** | Widespread in biobanks |
| **GOLD** | Environmental, biological & geographic metadata embedded | Metagenomics/environmental DNA | GOLD-specific | Internal GOLD portal | Extensive (>180 000 samples) |
| **ISBT 128** | Donation ID, product code, collection date, etc. | Clinical transfusion & Hospital Blood Management | International standard | Vendor/LIMS support | Global regulatory compliance |
| **Schrade et al.** | 17-char: taxonomy, sample type code, serial number | Wildlife/conservation biology | Prototype, project-specific | Registry proposed (unofficial) | Niche (wildlife studies) |
| **Kurtz et al.** | Donor, cell/tissue type, reprogramming method | Pluripotent stem cells | Published naming scheme | Manual implementation | Focused (stem-cell registries) |

\*\* SPRECware was proposed as an IT architecture/vision for integrations; SPRECbase was the initial concrete app released (23).

The TCGA barcode system provides hierarchical identifiers for biospecimens, enabling traceability from subject to aliquot. TCGA barcodes were programmatically generated by the Biospecimen Core Resource (BCR) using standardized rules within a Laboratory Information Management System (LIMS) system. Each barcode begins with a subject ID composed of the project name (“TCGA”), a tissue source site (TSS) code, and a unique subject number (e.g., ‘TCGA-02-0001’). This prefix is shared across all biospecimens from the same individual, making it easy to retrieve samples from a given donor. Subsequent segments encode specimen type (e.g., primary tumor), vial, portion, analyte (e.g., DNA/RNA), plate, and aliquot (e.g., ‘TCGA-02-0001-01C-01D-0182-06’). This comprehensive structure supports granular lineage tracing, such as tumor–normal comparisons, and the TCGA community has developed tools to facilitate its interpretation (22). Although the TCGA barcode served as the project’s primary identifier before the switch to UUIDs, its rigid, cancer-centric structure is confined to TCGA (and related GDC) studies and is not meant to be universal. ClarID adopts the same core concept but generalizes it through a YAML-based codebook, allowing metadata fields to be defined flexibly across diverse datasets. For subjects (individuals), additional fields are included to capture richer contextual information, while biosample components are abstracted for broader applicability. Importantly, biosample identifiers retain the subject ID, preserving the ability to search and group samples by donor. Note that components from the GDC data dictionary (https://docs.gdc.cancer.gov/Data_Dictionary/), such as center codes or study codes, can be reused within ClarID in the “project” identifier.

GTEx sample IDs are structured identifiers assigned by the GTEx Data Coordinating Center to uniquely label and trace biospecimens from donors through tissue collection and molecular processing. Each ID follows a standardized format, GTEX-<SubjectID>-<TissueCode>-SM-<AliquotID> (e.g., ‘GTEX-1117F-0126’), where the subject ID identifies the donor, the tissue code denotes the sampled organ or subregion, SM indicates the sample (aliquot) level, and the aliquot ID encodes preparation and sequencing batch information. These identifiers are centrally assigned through controlled pipelines to ensure consistency and traceability across the dataset. Tissue codes are derived from UBERON ontology terms but require external lookup for interpretation. The subject ID prefix is shared across all biospecimens from the same individual, enabling straightforward retrieval of all samples linked to a donor. ClarID retains this functionality while extending it with semantically richer fields, offering both human-readability and machine-parsability through a centralized, customizable vocabulary.

The SPREC system, widely used in biobanking for quality assurance, encodes preanalytical variables related to specimen collection, preparation, and storage in a fixed seven-element format. Supported by tools such as SPRECWare (23) and SPRECalc (24), both released in 2012, SPREC has since matured into version 4.0 (25), covering a broad range of biospecimen types and handling procedures. While the format follows a defined order of hyphen-separated elements, the meaning of each element differs between solid and fluid samples. For fluid samples, the elements describe sample type, primary container, pre- and post-centrifugation delays, centrifugation conditions, any second centrifugation, and long-term storage. For solid samples, they instead capture collection method, warm and cold ischemia times, fixation/stabilization method, fixation duration, and long-term storage. For example, the fluid sample code ‘PL1-SCI-A-C-M-H-A’ denotes a plasma sample single spun (PL1) collected in a sodium citrate tube (SCI), processed with a <2 h pre-centrifugation delay at room temperature (A), centrifuged 10–15 min at <3000 g and 2–10 °C (C), followed by a >48 h post-centrifugation delay at room temperature (M). Aliquoting was done within 1 h (H), and the sample stored at –85 to –60 °C (A). Whereas SPREC is designed for detailed biobank tracking and provenance, ClarID provides a flexible, open-source alternative that prioritizes encoding essential metadata fields for downstream analysis. Where appropriate, SPREC sample-type codes (covering fluids, solids, and cellular specimens) can also be leveraged within the ClarID “tissue” component.

The GOLD registry for metagenomics provides rich metadata for environmental and biological samples, but its identifiers are not semantically descriptive. ClarID addresses this by embedding key metadata directly into the identifier, enabling insight without reliance on external registries.

ISBT 128 is a globally standardized system (like ISBN from books) used for traceability of human biological products in clinical contexts. While highly regulated and comprehensive, it is not intended for research workflows. ClarID offers a non-regulatory alternative tailored for academic use cases that require interpretability and flexibility, without clinical compliance overhead.

Schrade et al. proposed a taxonomy-aware, 17-character identifier format for wildlife biospecimens that integrates species provenance and sample type. While informative, the system remains niche in both scope and adoption. ClarID builds on several of its core components. For example, it incorporates Element 2 from Component I (the first three letters of the genus plus the first three letters of the species, e.g., MacMul for *Macaca mulatta*) as species, whereas Element 1 (3-letter taxonomic code) is stored it in the codebook as “tax_code” (this element is not displayed by default in the ClarID identifier, it is retained internally for reference and disambiguation). Additionally, ClarID compresses the 6-character binomial into a 2-character representation in stub format using a Base62 transformation, based on the order of the species in the codebook. Overall, ClarID expands on the original concept with broader applicability, formal tooling, and support for both human-readable and compact representations.

Finally, Kurtz et al. designed a naming convention for pluripotent stem cell lines to ensure traceability across repositories. While well-structured, its use is limited to that specific domain. ClarID, in contrast, supports general use across organisms, assays, and experimental designs.

In summary, while these systems serve critical roles within their respective niches, ClarID seeks to fill the middle ground: offering a lightweight, interpretable, and reusable approach for embedding structured metadata directly into identifiers that are portable across datasets and research domains.

### Relation to Other Identifier Systems

Many biomedical repositories have developed identifier systems to ensure uniqueness and data organization. However, these identifiers typically lack embedded context, making them difficult to interpret without referencing external metadata.

For example, the International Cancer Genome Consortium (ICGC) assigns structured identifiers to donors (e.g., DO123456), specimens (SP654321), and aliquots (SA789012). While the prefixes indicate entity type, they contain no information about tissue, diagnosis, or assay. Similarly, platforms such as ENCODE, Roadmap Epigenomics, GEO, and ArrayExpress use accession numbers (e.g., ENCSR000AAA, GSM123456) that function purely as registry keys.

Although these identifiers ensure traceability, they introduce friction: researchers must rely on separate metadata files to understand sample context, often consulting additional documentation. This extra step can slow exploratory analysis and hinder integration into downstream workflows.

### Limitations and considerations

ClarID is not intended to replace primary identifiers such as UUIDs. Instead, it serves as a metadata-aware alternative that conveys key contextual information at a glance. In fact, multiple biosamples may share a ClarID when they come from the same subject under identical conditions, if batch or replicate information is not specified. One of the reasons TCGA transitioned from barcodes to UUIDs was that any change in metadata required updating the barcode. The same limitation applies to ClarIDs—when key metadata changes, the identifier may need to be regenerated.

One practical consideration concerns identifier length. The full-format ClarID (See Table 1) is semantically rich but may exceed length limits imposed by file systems, databases, or labeling hardware. To address this, a compact “stub” format is provided for use in constrained environments. The stub version retains part of the structure using abbreviated codes but is less interpretable without decoding. While many fields are shortened automatically, free-text fields like “project” require users to define a “stub_code” in the codebook for compact representation.

ClarID deliberately avoids encoding fine-grained technical or procedural details, such as sequencing platform, reagent lot, plate or vial identifiers, portion numbers or processing center. These are better managed in external metadata systems or LIMS, which can link to ClarID entries via UUIDs or internal sample IDs. The intent is to capture only the core biological and experimental context that is broadly useful for searching, filtering, tracking, and interpretation, rather than replicating full metadata registries. Additionally, if many fields are missing (e.g., marked as ‘NA’), the resulting identifiers may lose descriptive value and uniqueness, making them less useful for interpreting those entities.

ClarID includes a general-purpose codebook that supports many biomedical use cases, but it is not intended to be universally comprehensive. Users can adapt or extend the codebook by adding domain-specific terms, for example, custom assay types or experimental timepoints. ClarID-Tools supports this flexibility through example scripts that help align raw metadata into the required structure for batch encoding. This balance of centralized structure and local customization enables ClarID to support both harmonized and project-specific workflows. We hope this approach proves valuable to the community and encourages the development of diverse codebooks for specialized applications.

### Future directions

Looking ahead, we intend to apply ClarID across multiple European research projects in which we are actively involved. In parallel, we are assessing the integration of ClarID within the LIMS at CNAG to enable automated generation of structured identifiers during routine sample intake and processing. If external researchers adopt ClarID, a useful next step would be to establish a shared repository where users can find or contribute codebooks suited to their specific needs. Finally, we developed ClarID codes for the “subject” and “biosample” entities to address our immediate needs. However, the same design principles can be readily applied to other metadata-rich entities, or even files of any type, where self-descriptive identifiers would add value.

## CONCLUSIONS

ClarID introduces a practical, domain-agnostic framework for subject and biospecimen identification by embedding essential metadata, such as disease status, specimen type, and contextual attributes, into compact, structured identifiers. In contrast to project-specific systems such as TCGA or GTEx, ClarID is designed for broad applicability across biomedical and life science domains, supporting both human interpretability and machine-actionable parsing.

The specification is supported by an open-source implementation (ClarID-Tools) and a YAML-based codebook that defines controlled vocabularies. While it encourages the use of shared terms to foster semantic interoperability, it also supports local customization through configurable codebooks, allowing adaptation to project-specific needs. ClarID has been tested at scale on real-world datasets, including the GDC repository, and has the potential to be integrated into a variety of environments, such as LIMS platforms, analysis pipelines, and institutional databases.

Although not intended to replace full metadata barcoding systems, ClarID offers a lightweight and extensible solution to improve metadata search and accessibility, streamline downstream analysis, and promote better semantic integration. We encourage the research community to explore, adapt, and extend ClarID where concise, interpretable identifiers can add practical value.

## Data Availability

A copy of the downloaded input data, together with all processed results and scripts, is available at https://github.com/CNAG-Biomedical-Informatics/clarid-tools and can be reproduced via the linked Google Colab.

## AVAILABILITY AND REQUIREMENTS

- **Project name**: ClarID-Tools
- **Project home page**: https://github.com/CNAG-Biomedical-Informatics/clarid-tools
- **Operating system(s):** Linux, MacOS
- **Programming languages:** Perl 5 (ClarID-Tools), Python 3 (pre-processing scripts)
- **Other requirements:** Docker
- **License:** Artistic License 2
- **Any restrictions to use by non-academics**: None

## DOWNLOAD AND INSTALLATION

ClarID-Tools distribution is intended to run on any platform supported by Perl 5. We offer 3 installation options, including from: CPAN, containerized option (Docker), and GitHub source.

## SUPPLEMENTARY INFORMATION

**Additional file 1 (PDF):** Implementation details.

**Additional file 2 (PDF):** Supplementary table.

Table ST1: Implementation challenges and solutions.

**Additional file 3 (PDF):** Supplementary figures.

Figure SF1: Example QR codes of ClarID identifiers.

## DECLARATIONS

### ETHICS APPROVAL AND CONSENT TO PARTICIPATE

Not applicable.

### CONSENT FOR PUBLICATION

Not applicable.

### AVAILABILITY OF DATA AND MATERIALS

This study used publicly available data generated by multiple projects within the Genomic Data Commons (GDC) (https://portal.gdc.cancer.gov/) including but not limited to The Cancer Genome Atlas (TCGA) (https://www.cancer.gov/ccg/research/genome-sequencing/tcga). A copy of the downloaded input data, together with all processed results and scripts, is available at https://github.com/CNAG-Biomedical-Informatics/clarid-tools and can be reproduced via the linked Google Colab.

### AUTHORS’ CONTRIBUTIONS

MR conceived the study, developed ClariD-Tools, and wrote the manuscript. IGG obtained funding and revised the manuscript. All authors read and approved the final manuscript.

## ACKNOWLEDGEMENTS

We would like to thank Miranda Stobbe for her valuable suggestions and Ivo Leist for proofreading the manuscript. ChatGPT-4o by OpenAI was used to improve readability and language of some of the paragraphs.

## AUTHORS’ INFORMATION

## Authors and Affiliations

*Centro Nacional de Análisis Genómico, C/Baldiri Reixac 4, 08028 Barcelona, Spain*.

Ivo G. Gut, Manuel Rueda

*Universitat de Barcelona (UB), Barcelona, Spain*.

Ivo G. Gut, Manuel Rueda

