## Additional File 1 for "ClarID: A Human-Readable and Compact Identifier Specification for Biomedical Metadata Integration"

### Implementation details

#### Software

ClarID-Tools is implemented as a command-line interface (CLI) in Perl 5, selected for its efficiency in handling structured text files. The modular architecture and included test suite ensure that re-implementation in other programming languages is straightforward, particularly with support from modern LLMs.

Internally, the tool uses *Moo* and *MooX::Options* modules to define required and optional parameters, apply validation logic, and enable context-sensitive behavior depending on entity type and encoding mode. For data handling, it integrates *YAML::XS* and *Text::CSV_XS* for high-performance parsing of codebooks and bulk tabular files, along with *JSON::Validator* for schema-based validation of the codebook structure. QR code generation is performed using the *qrencode* library for Linux.

During development, we explored fully externalizing the ClarID specification itself, i.e., the structural rules governing identifiers, such as field widths, regex patterns, and transformations used for stub encoding (e.g., Base62). This was tested using JSON Schema. While attractive in theory, this approach quickly became impractical: managing the full specification through schema files led to greater complexity than the core implementation, with convoluted regex layers and transformations. Instead, we adopted a hybrid approach: some parameters (e.g., species, tissues, assays) remain configurable via the YAML codebook, while core structural rules are hardcoded for simplicity. The codebook itself is still validated against a JSON Schema to ensure consistency.

Although the current implementation focuses on identifiers for "subject" and "biosample" entities, the same principles (structured fields, consistent parsing rules, and YAML-based configuration) can be extended to additional entities such as cohorts, datasets, or experiments. As a temporary workaround (useful when code changes are not feasible) users may repurpose existing fields in the codebook for project-specific needs (e.g., reusing the "tissue" field to represent geographic location). While this departs from semantic accuracy, identifiers remain fully functional as long as the overall structure is preserved. With minor code modifications, such extensions can be supported natively.

**Encoding and decoding logic (stub format)**

In the human-readable form, identifiers are generated by concatenating fields with hyphens, with the only transformation being the zero-padding of the numeric "subject_id" field. The stub form, by contrast, applies compact transformations such as Base62 encoding for numeric fields, making it shorter but less directly interpretable. These transformations are described in detail below.

<u>Encoding of "project" (subject) and "study" (biosample)</u>

Labels such as 'TCGA_AML' are arbitrary alphanumeric strings rather than simple numbers, so there is no generic way to compress them into a compact stub. If a shorter code is desired, users should define a corresponding alias in the YAML codebook; otherwise, ClarID will include the full label in both human-readable and stub formats.

Base62 transformation of "subject_id" (subject and biosample)

In ClarID, each subject's numeric identifier ("subject_id") is converted into a fixed-width alphanumeric stub using Base62 encoding with a 62-character alphabet (0–9, A–Z, a–z). By default, the width is set to three characters, configurable via the --*subject_id_pad_lengt*h flag.

For "subject_id" = 0, the stub is simply "0" repeated to the chosen width. For values greater than zero, successive division and modulo operations by 62 are performed to extract characters, which are then left-padded with "0" to enforce the fixed width. For example, subject_id = 999 yields "G7," which is padded to "0G7."

Although Base62 encoding naturally produces variable-length strings, enforcing a fixed width ensures consistent parsing and supports up to 62^3 – 1 = 238,327 unique individuals with a three-character stub. Larger cohorts can be accommodated by increasing the width using the --*subject_id_base62_width* flag.

Encoding of "condition" (subject and biosample)

The "condition" field is encoded by mapping each ICD-10 code to a corresponding numerical index using a predefined internal JSON file. That number is then converted to a Base62 representation using the same transformation method as for "subject_id". The resulting alphanumeric code is fixed to a length of 3 characters to maintain compactness. This allows disease information to be stored concisely while still allowing for reverse mapping during decoding. This field supports multiple values: in the human-readable format they are separated by the '+' character, while in stub form they are concatenated using their stub codes.

Encoding of "species" (biosample)

Species "stub_code" is declared statically in the codebook and represented using two-character codes, again based on the 62-symbol alphabet (0–9, A–Z, a–z). This allows encoding of up to 3,844 distinct values (with one code, such as "00," reserved for unknown species). Although the codebook includes an optional taxonomy-based identifier (tax_code, e.g., "MacMul" for *Macaca mulatta*) for traceability, this is not used in the stub format.

Encoding/decoding of "tissue", "sample_type" and "assay" (biosample)

These fields are expected to have a limited number of distinct values (dozens to hundreds), so instead of using dynamic base encoding, we adopted predefined stub_codes. This choice provides greater flexibility and interpretability while keeping stub strings short.

During decoding, the parser first handles fixed-width fields (e.g., subject_id) and then applies greedy reverse lookup for the variable-length fields. Stub codes are sorted in descending order of length to prevent prefix collisions. For example, with stub codes such as "PB" (peripheral blood), "T" (tumor), and "HI" (Hi-C sequencing), a stub like …PBTHI… will be correctly parsed into its respective metadata components. Although there is no strict length limit for stub codes, we recommend codes between 2 and 5 characters to maintain compactness and readability.

**Raw data pre-processing script**

A custom Python 3 pre-processing script reads raw TSV/CSV data, applies YAML-defined field-by-field transformations (e.g., trimming, value mapping, multi-value normalization, etc.), producing a standardized CSV ready for ClarID-Tools.

Pre-processing of timepoint's "duration" (biosample)

To keep the component to 3 characters (human format) or 2 (stub format), the pre-processing script converts a numeric day count into the smallest allowed ISO-8601 bin (P[0-9][DWMY]) that can represent it without exceeding 9 in that unit, escalating from days → weeks → months → years, with rounding as configured.

**Codebook (v0.02)**

The codebook and ClarID-Tools are versioned synchronously, and the latest release is available at the project repository. The full codebook is too lengthy to include here; an example is provided.

```
metadata:
  version: "0.02"
  local_version: "CNAG-2025.09.05" # optional
  author: "Manuel Rueda"
  center: "CNAG"
  date: "2025-09-05"
  description: "CNAG's ClarID codebook."
  repository: "https://github.com/CNAG-Biomedical-Informatics/clarid-tools"

entities:
  biosample:
    project:
      TCGA-AML:
        code: TCGA_AML
        stub_code: AML
        label: "TCGA Acute Myeloid Leukemia"
        id: "NCIT:C17998"
```

```
    species:
      Human:
        code: HomSap
        stub_code: "01"
        label: "Homo sapiens"
        id: "NCBITaxon:9606"
        tax_code: MPH
    tissue:
      Liver:
        code: LIV
        stub_code: L
        label: "Liver"
        id: "UBERON:0002107"
    sample_type:
      Tumor:
        code: TUM
        stub_code: T
        label: "Neoplasm"
        id: "NCIT:C3262"
    assay:
      RNA_seq:
        code: RNA
        stub_code: R
        label: "RNA-seq"
        id: " EFO:0008896"
    timepoint:
      Baseline:
        code: BSL
        stub_code: "B"
        label: "Baseline"
        id: "NCIT:C25213"
    condition_pattern:
      regex: "^([A-Z]\\d{2}(?:\\.\\d+)?)$"
      code_format: "%s"
      stub_format: "%s"
    duration_pattern:
            regex: '^(?:P?(\d+)([DWMY])|P?(0)(N))$'
      code_format: "P%d%s"
      stub_format: "%d%s"
    batch_pattern:
      regex: "^(\\d{1,2})$"
      code_format: "B%02d"
      stub_format: "%02d"
    replicate_pattern:
      regex: "^(\\d{1,2})$"
      code_format: "R%02d"
      stub_format: "%02d"

  subject:
    study:
      TCGA-AML:
        code: TCGA_AML
        stub_code: AML
        label: "TCGA Acute Myeloid Leukemia"
        id: "NCIT:C17998"
```

```
type:
  Case:
    code: Case
    stub_code: C
    label: "Case Study"
    id: " NCIT:C15362"
sex:
  Male:
    code: Male
    stub_code: M
    label: "Male"
    id: "PATO:0000384"
age_group:
  Age20to29:
    code: A20_29
    stub_code: A2
    label: "Age 20-29"
    id: "NCIT:C20192"
```


**Specification (v0.02)**

The specification and ClarID-Tools are versioned synchronously. Please use this link to get the latest version.

### Biosample - Human format

**Delimiter:** -

| # | Component | Source field | Type | Pattern / Format | Built from |
|---|---|---|---|---|---|
| 1 | Project | project.code | string | free string | codebook value (if present) |
| 2 | Species | species.code | string | 6-letter binomial acronym (e.g. HomSap) | codebook value |
| 3 | Subject ID | subject_id | integer -> str | zero-pad to 5 digits (default) | sprintf("%0${pad}d", $sid) |
| 4 | Tissue | tissue.code | string | exactly 3 letters [A-Z]{3} | codebook value |
| 5 | Sample Type | sample_type.code | string | exactly 3 letters [A-Z]{3} | codebook value |
| 6 | Assay | assay.code | string | exactly 3 letters [A-Z]{3} | codebook value |
| 7 | Condition | condition | string | ICD-10 diagnose | used verbatim (or |

| | | | | | |
|---|---|---|---|---|---|
| | | | | code(s) [A-Z]\d{2}(?:\.\d+)? (<=10) | concatenated with +) |
| 8 | Timepoint | timepoint | string | alphanumeric events e.g. Baseline | codebook value |
| 9 | Duration | duration | string | ISO 8601 3-char (P1D,P7W,P3M,P1Y) or P0N (Not Available) | duration_pattern |
| 10 | Batch (opt) | batch | integer -> str | B%02d (e.g. B01) | batch_pattern |
| 11 | Replicate (opt) | replicate | integer -> str | R%02d (e.g. R05) | replicate_pattern |

### Biosample - Stub format

**Delimiter:** (none)

| # | Component | Source field | Type | Pattern / Format | Built from |
|---|---|---|---|---|---|
| 1 | Project stub | project.stub_code | string | free string | codebook value |
| 2 | Species stub | species.stub_code | string | 2-char codebook stub (Base62 alphabet) | codebook value |
| 3 | Subject stub | subject_id | integer -> Base62 | width 3 (default) - max 238,327 | 3-char Base62 from integer |
| 4 | Tissue stub | tissue.stub_code | string | 1-3 chars | codebook value |
| 5 | Sample Type stub | sample_type.stub_code | string | 1-3 chars | codebook value |
| 6 | Assay stub | assay.stub_code | string | 1-3 chars | codebook value |
| 7 | Condition stub | condition | code -> Base62 | N x 3-char Base62 stubs + 2-digit count (%02d) | codebook order + 3-char Base62 from integer |
| 8 | Timepoint stub | timepoint.stub_code | string | 1-2 chars | codebook value |

| | | | | | |
|---|---|---|---|---|---|
| 9 | Duration stub | duration | string | digits+unit (e.g. 7W) | duration_pattern |
| 10 | Batch stub (opt) | batch | integer | B%02d (e.g. B01) | batch_pattern |
| 11 | Replicate stub (opt) | replicate | integer | R%02d (e.g. R05) | replicate_pattern |

### Subject - Human format

**Delimiter:** -

| # | Component | Source field | Type | Pattern / Format | Built from |
|---|---|---|---|---|---|
| 1 | Study | study | string | free string | codebook value (if present) |
| 2 | Subject ID | subject_id | integer -> str | zero-pad to 5 digits (default) | sprintf("%0${pad}d",$sid) |
| 3 | Type | type.code | string | codebook codes | codebook value |
| 4 | Condition | condition | string | ICD-10 diagnose code(s): [A-Z]\d{2}(?:\.\d+)? (<=10) | used verbatim (or concatenated +) |
| 5 | Sex | sex.code | string | codebook codes | codebook value |
| 6 | Age Group | age_group.code | string | codebook codes | codebook value |

### Subject - Stub format

**Delimiter:** (none)

| # | Component | Source field | Type | Pattern / Format | Built from |
|---|---|---|---|---|---|
| 1 | Study stub | study.stub_code | string | free string | codebook value (if present) |
| 2 | Subject ID stub | subject_id | integer -> Base62 | width 3 (default) - max 238,327 | 3-char Base62 from integer |
| 3 | Type stub | type.stub_code | string | 1 char | codebook |

| | | | | | |
|---|---|---|---|---|---|
| | | | | | value |
| 4 | Condition stub | condition | code -> Base62 | N x 3-char Base62 stubs + 2-digit count (%02d) | codebook order + 3-char Base62 from integer |
| 5 | Sex stub | sex.stub_code | string | 1 char | codebook value |
| 6 | Age Group stub | age_group.stub_code | string | 2 chars | codebook value |
