## Additional File 2 for "ClarID: A Human-Readable and Compact Identifier Specification for Biomedical Metadata Integration"

### Supporting Table 1: Implementation challenges and solutions

| Data pre-processing | |
|---|---|
| **Faced challenges** | **Solution** |
| CSV is flexible but unstructured; missing values and inconsistent codes are common | We developed a preprocessing script, guided by a configuration file, to standardize raw CSV rows and columns. The script handles missing values, harmonizes inconsistent codes, and produces a clean CSV suitable for bulk processing with ClarID-Tools. The scripts are available at: https://github.com/CNAG-Biomedical-Informatics/clarid-tools/tree/main/nb/data/scripts |
| **CLI + Module** | |
| **Faced challenges** | **Solution** |
| Standardization of components' vocabulary | In the codebook, ontology terms were used to define properties where possible (e.g., tissues with UBERON). For diseases and technical fields like batch/replicate, regex rules were applied. The structure remains flexible, allowing users to extend it as needed. |
| Externalized schema vs. hard-coded specification | Full JSON Schema externalization proved impractical; we adopted a hybrid approach with YAML-configurable parameters and some hardcoded elements, while validating the codebook itself against a JSON Schema. |
| Compaction of human → stub formats | Certain fields required Base62 encoding to condense information. The field width was restricted for parsing but can be adjusted through CLI parameters. |
