## Additional File 3 for "ClarID: A Human-Readable and Compact Identifier Specification for Biomedical Metadata Integration"

### Supporting Figure 1

**(a)**

AsthmaCohort-01002-Control-J98.51-Female-A50_59

AsthmaCohort-01004-Case-J98.51-Male-A40_49

COPDStudy-01001-Case-J98.51-Male-A40_49

COPDStudy-01003-Control-J98.51-Female-A50_59

**(b)**

CNAG_Test-DanRer-00003-BLO-NOR-WES-I46-SUR-P1M-B03-R01

CNAG_Test-HomSap-00001-LIV-NOR-RNA-C22.0-BSL-P0D-B01-R05

CNAG_Test-MusMus-00002-BRN-TUM-CHI-C71.0-TRT-P7W-B02-R02

CNAG_Test-RatNor-00004-KID-NOR-LCMS-C66-CHL-P3Y-B01-R10

**Figure SF1.** Example QR (quick-response) codes with their decoded ClarID strings shown above each image, for (a) a "subject" entity and (b) a "biosample" entity. The codes can be read by any QR-enabled device, such as a smartphone.
